# In-utero exposure to PM_2.5_ and adverse birth outcomes in India: Geostatistical modelling using remote sensing and demographic health survey data 2019-21

**DOI:** 10.1101/2024.09.16.24313773

**Authors:** Arup Jana, Malay Pramanik, Arabinda Maiti, Aparajita Chattopadhyay, Mary Abed Al Ahad

## Abstract

Rapid urbanization in India is contributing to heightened poor air quality. Yet research on the impact of poor air on adverse birth outcomes (ABOs) especially in the public health aspect is less in India. This study investigates the influence of air quality on birth weight (LBW) and preterm birth (PTB). Utilizing data from the National Family Health Survey and satellite images, the study employs various statistical analyses and spatial models to elucidate the connection between in-utero exposure to air pollution and birth outcomes, both at the individual and district levels. It was observed that approximately 13% of children were born prematurely, and 17% were born with low birth weight. Increased ambient PM_2.5_ concentrations during pregnancy were associated with higher odds of LBW (AOR: 1.4; 95% CI: 1.29–1.45). Mothers exposed to PM_2.5_ during pregnancy had a heightened likelihood of delivering prematurely (AOR: 1.7; 95% CI: 1.57–1.77) in comparison to unexposed mothers. Climatic factors such as rainfall and temperature had a greater association with ABOs. Children residing in the Northern districts of India appeared to be more susceptible to the adverse effects of ambient air pollution. Furthermore, indoor air pollution was found to be associated with LBW. Employing a distributed spline approach, the study identified a discernible upward trend in the risk of adverse birth outcomes as the level of exposure increased, particularly following an exposure level of 40 PM_2.5_ ug/m^3^. Among the spatial models employed, the MGWR spatial model exhibited the highest level of goodness of fit. In addition to addressing immediate determinants such as nutrition and maternal healthcare, it is imperative to collaboratively address distal factors encompassing both indoor and outdoor pollution to attain lasting enhancements in child health.

**Highlights:**

- An estimated 13% of the children were preterm and 17% were low birth weight in India in 2019-21.
- High PM_2.5_ concentration during pregnancy was associated with LBW and PTB.
- Children residing in the Northern districts of India were found to be more vulnerable to poor air quality.
- A rising trend in the risk of adverse birth outcomes was observed after exposure level of 40 PM_2.5_ ug/m3.
- The MGWR spatial model demonstrated the highest level of goodness of fit.

**Graphical abstract:** 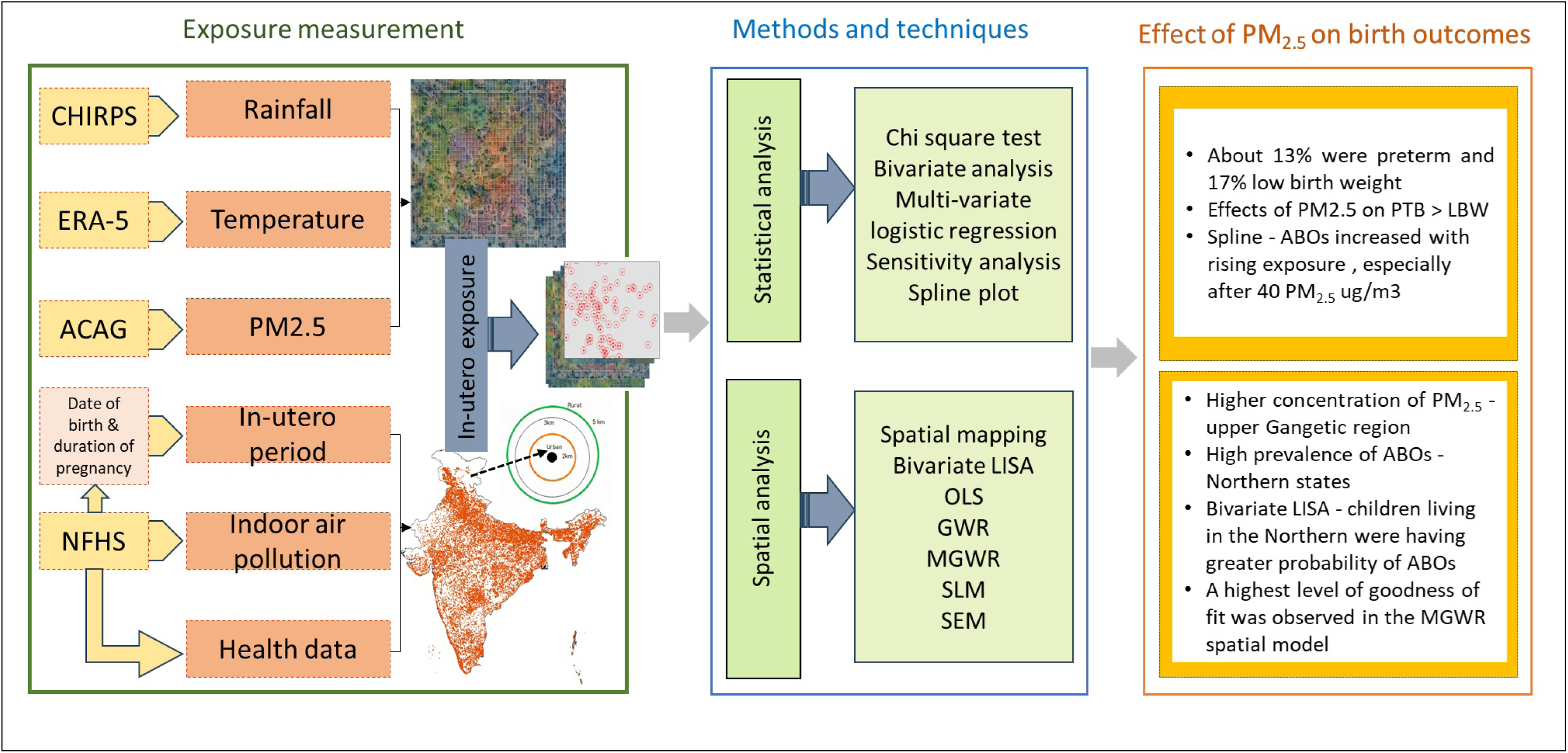

## 1. Introduction

Ambient air pollution poses an existential global environmental threat to planetary and human health, with a disproportionate burden of its detrimental effects falling on those residing in low and middle-income countries ^1^. Consequently, the United Nations Climate Change Conference has urged developed countries to provide financial support to less developed and developing countries to confront the adverse impacts of air pollution and establish appropriate mechanisms to mitigate climate change ^2^. Besides being a significant driver of climate change, air pollution is the most critical risk factor for adverse health consequences ^3^. Referred to as the "silent killer," ambient air pollution is among the top five risk factors for mortality in both males and females ^4–6^. In 2019 alone, ambient particulate matter pollution was responsible for 118 million disability-adjusted life years (DALY) and 4.14 million deaths in 2019 ^7^. Among air pollutants, ambient fine particulate matter (PM_2.5_) is considered the most harmful air pollutant [4, 8–10]. These particles primarily originate from the burning of fossil fuels and biomass ^11^. With a diameter of less than 2.5 microns, exposure to these particles increases the risk of respiratory diseases, lung cancer, stroke, and heart disease ^9,12–16^.

In the 2023 World Air Quality Report, India was ranked as the third most polluted country out of 134 nations based on its average yearly PM2.5 levels ^17^. Notably, about 7 out of 10 Indians are exposed to air pollution levels that exceed the national standard of 40 µg/m3 ^18^. Alarmingly, in 2019, as many as 0.98 million deaths in India were attributed to ambient particulate matter pollution ^19^. The key factors of this alarming trend are primarily attributed to the rising air pollution levels in the country due to urbanization and industrialization ^1,20,21^. To address this critical air pollution situation, the government of India introduced the National Clean Air Program in 2019, setting a targeted reduction in air pollution. It envisages reaching a minimum 20% reduction in particulate matter concentration by 2024, compared to 2014 levels ^22,23^. However, despite these significant efforts, India still ranks as the second most polluted country in the world ^22^.

While air pollution has adverse impacts on the health of individuals across all age groups, infants and children are considered more vulnerable due to their developing organs and higher air intake per unit of body weight ^24,25^. Ambient air pollution has been associated with a range of pediatric morbidities, including adverse birth outcomes (ABO), asthma, cancer, and an increased risk of chronic diseases in life stages ^9,13,26–30^. Among the ABOs associated with ambient air pollution are preterm birth (PTB) and low birth weight (LBW) ^25,27,31,32^. PTB and LBW are important predictors of under-five mortality and malnutrition ^33,34^. The World Health Organization estimates that approximately 15 million babies are born preterm (<37 weeks) each year ^34^. Furthermore, 15% of babies worldwide are born with LBW, defined as <2500 grams at birth ^35^. While LBW is a global public health concern, its prevalence is particularly higher in low and middle-income countries ^36^.

A systematic review of 41 studies found that exposure to PM_2.5_ is associated with PTB, LBW, and small-for-gestational-age births ^37^. A prospective cohort study conducted in Tamil Nadu provided the first quantitative evidence linking rural-urban PM_2.5_ exposures during pregnancy of LBW ^38^. This study concluded that increased exposure to particulate matter exposure during pregnancy is associated with a decrease in birth weight among newborns ^39^. However, research in developing countries, albeit limited, established that maternal exposure to ambient air pollution is associated with higher odds of LBW and PTB among children ^40–43^.

The majority of studies investigating the association between ambient air pollution with ABOs have primarily been conducted in high-income countries ^36,40^. The evidence regarding this particular issue is somewhat less in developing countries ^32^. According to National Family Health Survey-5, 18% of children born in the five years preceding the survey had low birth weight. India has been identified as a significant contributor to global preterm births ^44^. Moreover, ambient air pollution levels are expected to increase in India as the country is transitioning from a rural economy to an urban one, with the urban population projected to reach 53% by 2050 from 35.87 in 2022 ^45^. Despite the alarming rise in air pollution levels in India, there has been a paucity of research exploring its impact on ABOs. Moreover, the findings of a spatial analysis on the effects of air pollution on adverse birth outcomes (ABOs) will be helpful for implementing area-specific schemes for policymakers or stakeholders, which has not yet been done at the national level in India. Therefore, the study hypothesizes a negative impact of PM_2.5_ on birth outcomes in India. In order to address this research gap in the available evidence, the present study endeavors to investigate the impact of ambient air pollution on ABOs, particularly focusing on LBW and PTB at the national level. Further, the study utilized different geospatial models to highlight the vulnerable areas that need to be focused on, with robust estimated outcomes controlling for environmental, demographic, and socioeconomic confounding factors. The study utilizes nationally representative data from the latest round of the NFHS, 2019-21, thus ensuring the generalizability of results ^46^. The results of this study might be imperative as LBW and PTB continue to be significant public health concerns in India. Moreover, this study will augment the existing but limited literature focused on investigating the association of ambient air pollution with LBW and PTBs, a question of paramount public health significance.

## 2. Data and Methods

### 2.1. Population and health data

Population data were extracted from the fifth NFHS, conducted across 36 states and union territories at the national level. A stratified two-stage sampling procedure was employed in the survey design. Primary Sampling Units (PSUs) were villages in rural areas and census enumeration blocks (CEBs) in urban areas. Survey questionnaires were prepared to take out detailed information on maternal and child health, as well as socioeconomic data. Additionally, the NFHS data incorporated the collection of biomarker data ^46^. The survey encompassed reproductive history, including data on contraceptive use, pregnancy, birth, termination, and abortion. Children born 0 to 5 years preceding the survey were considered in the research. The protocol for the NFHS-5 survey, including the content of all the survey questionnaires, received approval from both the International Institute of Population Sciences (IIPS) institutional review board and the ICF institutional review board. Furthermore, it underwent review protocol by the U.S. Centres for Disease Control and Prevention (CDC). Further details on the sampling design can be obtained in the Indian National Report (https://dhsprogram.com/pubs/).

### 2.2. Air pollution data

In this study, global level PM_2.5_ data was derived from the Atmospheric Composition Analysis Group (ACAG), to measure the exposure of mothers during the pregnancy period ^47^. The spatial resolution of PM_2.5_ was 0.01° × 0.01° and Resolution-Tiered Approach (RTA) was employed to estimate the concertation of PM_2.5._ The data is prepared by the fusion of multiple sensors including Aerosol Optical Depth (AOD) measurements from NASA’s Moderate–Resolution Imaging Spectro-radiometers (MODIS), Multi-Resolution Imaging Spectro-Radiometers (MISR), and Sea–viewing Wide Field–of–View Sensor (SeaWiFS) instruments with the Goddard

Earth Observing System (GEOS) and chemical transport model (Chem) data. The data was further calibrated to match global ground-based observations using a Geographically Weighted Regression (GWR) technique, and its accuracy has been validated in several previous studies ^43,48–55^. Each survey participant was assigned an average in-utero air pollution exposure based on the child’s date of birth and the duration of pregnancy, as recorded in the NFHS survey data. For instance, if a child was born in January 2019 with a pregnancy duration of 9 months, the in-utero exposure period was considered from May 2018 to the end of January 2019. The cluster points were randomly displaced by 2 and 5 km in urban and rural areas, respectively, to maintain the privacy of participants. Thus, a 3 km buffer was created around each cluster point for the extraction of pollutant data.

### 2.3 Climate data

We used the Climate Hazards Group InfraRed Precipitation with Station (CHIRPS) precipitation dataset, covering a span of 43 years from 1981 to the present ^56^, with a spatial resolution of 5 km (https://www.chc.ucsb.edu/data/chirps). This high-quality dataset served as the foundation for developing a comprehensive and robust climatic index, laying the groundwork for deeper insights into the long-term trends and patterns of climatic hazards in various geographical areas.

We also utilized the ERA5-Land product, distributed by the Copernicus Climate Change Service (C3S) Climate Data Store (CDS) of ECMWF, to analyze land surface temperature (LST) in our study ^57^. This dataset provides monthly temperature values at a spatial resolution of 0.1 degrees (approximately 11,200 meters at the equator) on a latitude/longitude Climate Modelling Grid (CMG), covering the period from January 1981 onwards. The ERA5-Land product, along with its fine resolution and long-term availability, enables detailed assessments of LST variations and their implications for various ecosystems and human activities ^56,58–60^.

### 2.4 Variable description

#### 2.4.1. Dependent variables

According to the World Health Organization (WHO), Preterm birth is defined as a live birth before 37 completed weeks of gestation, while low birth weight is defined as weight at birth less than 2500 grams ^61^. In the present study, we followed the methodology outlined by Jana, Banerjee, and Khan (2023) ^62^, to measure preterm birth using the calendar method of the Demographic and Health Surveys (DHS). The NFHS collected data on birth weight using the following questions: Was (name of the child) weighed at birth? How much did (name of the child) weigh? The information was reported in two ways; first, the mother recalled about her baby’s weight, and second, reported with the help of any card of their baby’s weight ^46^. The other outcome variable in this study was preterm birth, which is estimated based on the duration of pregnancy. Both outcome variables are dichotomous, with ‘0’ signifying that the child did not have LBW/was not preterm, and ‘1’ indicating that the child had LBW/was preterm.

#### 2.4.2. Model covariates

We selected the factors associated with adverse birth outcomes based on published literature and the availability of variables within the dataset. The study included the child’s characteristics, such as the sex of the child (male and female) and birth order (1^st^, 2^nd^ and 3^rd^ & above), which were considered as confounding factors. Babies born at home was defined as non-institutional delivery and delivery taken place at any medical institution was considered as institutional delivery. Mother’s general health plays a pivotal role in determining the health of newborns ^63,64^. NFHS-5 collected anthropometric measurements using biomarkers. These measurements were used to calculate Body Mass Index (BMI) by dividing weight in kilograms by height in meters squared (kg/m^2^). BMI was categorized into four groups; thin (BMI <18.5), normal (BMI 18.5–24.9), overweight (BMI 25–30), and obese (BMI≥30.0).

The model incorporated several maternal variables, such as mother’s level of education (categorized as illiterate/primary, secondary, and higher education) and mother’s age at the time of childbirth (< 20, 20 – 24, 25-29, and 30 years & above). In the survey, respondents were asked about their primary cooking fuel source. We classified wood, coal, dung cake, and crop residue as solid or unclean fuel and natural gas, liquefied petroleum gas, and electricity as clean fuels. The household’s wealth quintile was categorized into three categories; ‘poor’, ‘middle’ and ‘rich’. Additionally, the household’s religion was recoded as ‘Hindu’, ‘Muslim’ and ‘Others’ (such as Sikh, Christian, Jain etc.).

### 2.5. Statistical analysis

The weighted prevalence of LBW and PTB was estimated using the exposed sample, and the Chi-square test (χ2) was performed to evaluate the association between dependent and independent variables. Geospatial analysis has been conducted using ArcGIS (version 10.8) to prepare the prevalence maps. Further, multivariate logistic regression was employed to examine the association between exposure to PM_2.5_ during pregnancy and birth outcomes after controlling all the possible confounding factors. The logistic regression model is defined as,

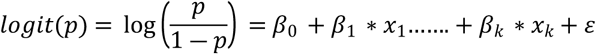

Where, *β*_0_ is intercept and β1… βk are regression coefficients indicating the relative effect of a particular explanatory variable on the outcome, while *ε* is an error term. Further, bivariate Local Indicators of Spatial Association (LISA) was used in the study to explore the spatial association between exposure to PM_2.5_ and adverse birth outcomes. A sensitivity analysis was employed by adjusting different determinants to explore the relation between in-utero exposure to PM_2.5_ and birth outcomes such as PTB and LBW. Further, in the study, a spline function was utilized to visually depict the connection. This connection involved the likelihood of occurrences such as having children with LBW and PTB, all influenced by exposure to air pollution during pregnancy. A spline function integrates several polynomial sections linked by knots, forming a continuous curve. The quantity of knots can be modified to control the curve and smoothness, increasing or decreasing it as needed. To determine knot placement, we employed quantiles of the exposure level of PM_2.5_ in the individuals.

Bivariate LISA measures the local correlation between a variable and the weighted average of another variable in the neighborhood.

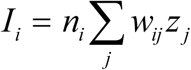

Where, Z_i_ denotes standardized variable of interest and W_ij_ is weight matrix. The bivariate LISA functionality the cluster maps of the association between two variables. The cluster map is a special choropleth map showing those locations with a significant local Moran statistic classified by the type of spatial correlation: bright red for high-high associations, bright blue for low-low, light blue for low-high and light red for high-low. The high-high and low-low suggest clustering of similar values, whereas high-low and low-high locations indicate spatial outliers. The scatter plot of the dependent and independent variables of a spatial unit shows the statistical figure of the association.

Several geostatistical models were used in this study to identify the best-fitting model, and the results will be helpful for policymakers to launch area-specific schemes for vulnerable regions… First, Ordinary Least Squares (OLS) technique was used. This is a classic linear regression model that seeks to find the best-fitting line through the data by minimizing the sum of squared differences between observed and predicted values. OLS assumes that the relationship between the dependent and independent variables is constant across space. Second, we employed Geographically Weighted Regression (GWR) which is a spatial regression technique that allows for the exploration of spatially varying relationships between variables. It recognizes that the relationships may change across different geographic locations, providing a localized understanding of the data. Finally, Multiscale Geographically Weighted Regression (MGWR) which extends GWR by considering relationships at multiple spatial scales was applied ^65^. This model accounts for varying relationships not only at the local level but also extends its analysis across different geographic scales, providing a more comprehensive analysis of spatial variability. The model employs two spatial regression models, namely the Spatial Lag Model (SLM) and the Spatial Error Model (SEM), each addressing distinct aspects of spatial dependency. The Spatial Lag Model (SLM) is a type of spatial autoregressive model that considers the spatial dependency of observations. It assumes that the values of the dependent variable are influenced by the values of neighboring observations, incorporating spatial relationships into the model ^66^. It accounts for the spatial interdependence of data points. On the other hand, the Spatial Error Model (SEM) is another spatial regression model that accounts for spatial autocorrelation in the error terms of the model. It assumes the presence of spatially correlated errors that are not captured by the independent variables, allowing for a more accurate representation of the data’s spatial structure.

## 3. Results

### 3.1 Characteristics of the sample

**Table 1** represents the percentage distribution of the sample size used in the present study. Approximately 52% of the sample consisted of female children, and the rest of the sample was male children. Among the sample, 13% of the sample were born preterm, whereas 17% of the sample were LBW. About half of the sample belonged to 2^nd^ birth order category. One out of 10 mothers were teenagers (under 20 years of age). About 14% of the children were born at home, and nearly 19% of the mothers were undernutrition or categorized as ‘thin’. Half of the sample belonged to households classified as ‘poor’, and 73% of the sample belonged to the Hindu religion. Additionally, 58% of the households reported using solid fuels for cooking.

**Table 1.** Sample distribution used in the analysis.

| **Variables** | **Sample** | **Percentage** |
| --- | --- | --- |
| **Preterm birth** |  |  |
| No | 2,01,657 | 87.45 |
| Yes | 28,933 | 12.55 |
| **Low birth weight** |  |  |
| No | 1,71,116 | 82.6 |
| Yes | 36,039 | 17.4 |
| **Sex of the child** |  |  |
| Male | 1,19,474 | 51.81 |
| Female | 1,11,116 | 48.19 |
| **Birth order** |  |  |
| 1 | 88,235 | 38.26 |
| 2 | 1,11,898 | 48.53 |
| 3+ | 30,457 | 13.21 |
| **Mother's age at delivery** |  |  |
| Below 20 | 26,282 | 11.4 |
| 20-24 | 98,137 | 42.56 |
| 25-29 | 68,494 | 29.7 |
| 30&above | 37,677 | 16.34 |
| **Place of delivery** |  |  |
| Home | 31,331 | 13.59 |
| Hospital | 1,99,259 | 86.41 |
| **Place of residence** |  |  |
| Rural | 1,84,038 | 79.81 |
| Urban | 46,552 | 20.19 |
| **Mother's body mass index** |  |  |
| Underweight | 42,112 | 18.73 |
| Normal | 1,42,504 | 63.38 |
| Overweight/obese | 40,208 | 17.88 |
| **Mother's education** |  |  |
| Illiterate/primary | 80,636 | 34.97 |
| Secondary | 1,18,602 | 51.43 |
| Higher | 31,352 | 13.6 |
| **Wealth status** |  |  |
| Poor | 1,16,744 | 50.63 |
| Middle | 44,700 | 19.39 |
| Rich | 69,146 | 29.99 |
| **Religion** |  |  |
| Hindu | 1,69,233 | 73.39 |
| Muslim | 33,194 | 14.4 |
| Others | 28,163 | 12.21 |
| **Cooking fuel** |  |  |
| Clean fuel | 97,582 | 42.32 |
| Solid fuel | 1,33,008 | 57.68 |

### 3.2 Spatial distribution of ABO and PM_2.5_

The spatial distribution of ambient PM_2.5_ shows a high concentration over the upper Gangetic region (**Figure 1**), covering states like Uttar Pradesh, Bihar, Delhi, Punjab and Haryana, whereas a lower concentration was observed in the Southern and North-Eastern regions of India. The highest prevalence of preterm was observed in the Northern states, such as Himachal Pradesh (39%), Uttarakhand (27%) and Rajasthan (18%), and Delhi (17%), including North-Eastern states like Nagaland (**Table S1**). In contrast, a lower prevalence of PTB was observed in Mizoram, Manipur and Tripura. As for LBW, the highest prevalence was in Punjab (22%), followed by Delhi, Dadra and Nagar Haveli, Madhya Pradesh, Haryana and Uttar Pradesh. Conversely, fewer children with LBW were observed in the North-East region of India (**Figures 1b and 1c**).

**Figure 1.**
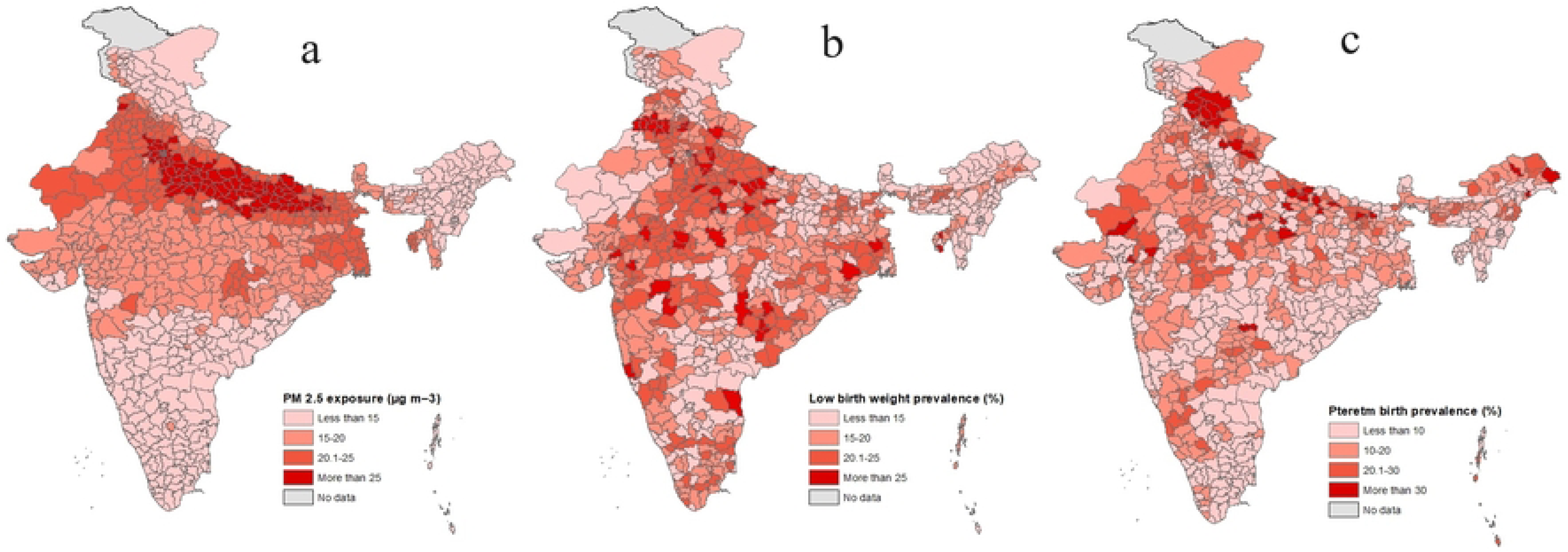
(a) Spatial distribution of in-utero exposure to PM_2.5,_ (b) low birth weight and (c) preterm birth across in India

### 3.3 Effects of PM_2.5_ on ABO at the individual level

Figure 2 shows the weighted percentage of LBW and PTB by selected background characteristics. The observation revealed that the prevalence of LBW was higher among females (20%) compared to males (17%), whereas a significant difference was not observed for PTB (**Table S2**). In both cases, the proportion was higher among teenage mothers. However, a decreasing percentage was found with age for LBW, but an increasing level of PTB can be observed after age 30 years. The percentage of adverse birth outcomes was greater among children born at home and residing in rural areas. Underweighted mothers had more LBW (22%) and PTB (13%) children than normal mothers, 17% and 12%, respectively. Illiterate and primary educated mothers were having more LBW and PTB children. A similar result can be observed for wealth status. Poor mothers had more adverse birth outcomes. However, mothers who belonged to the Muslim religion had a lower percentage of LBW and a higher proportion of PTB. Households using solid fuels for cooking had more adverse birth outcomes.

**Figure 2.**
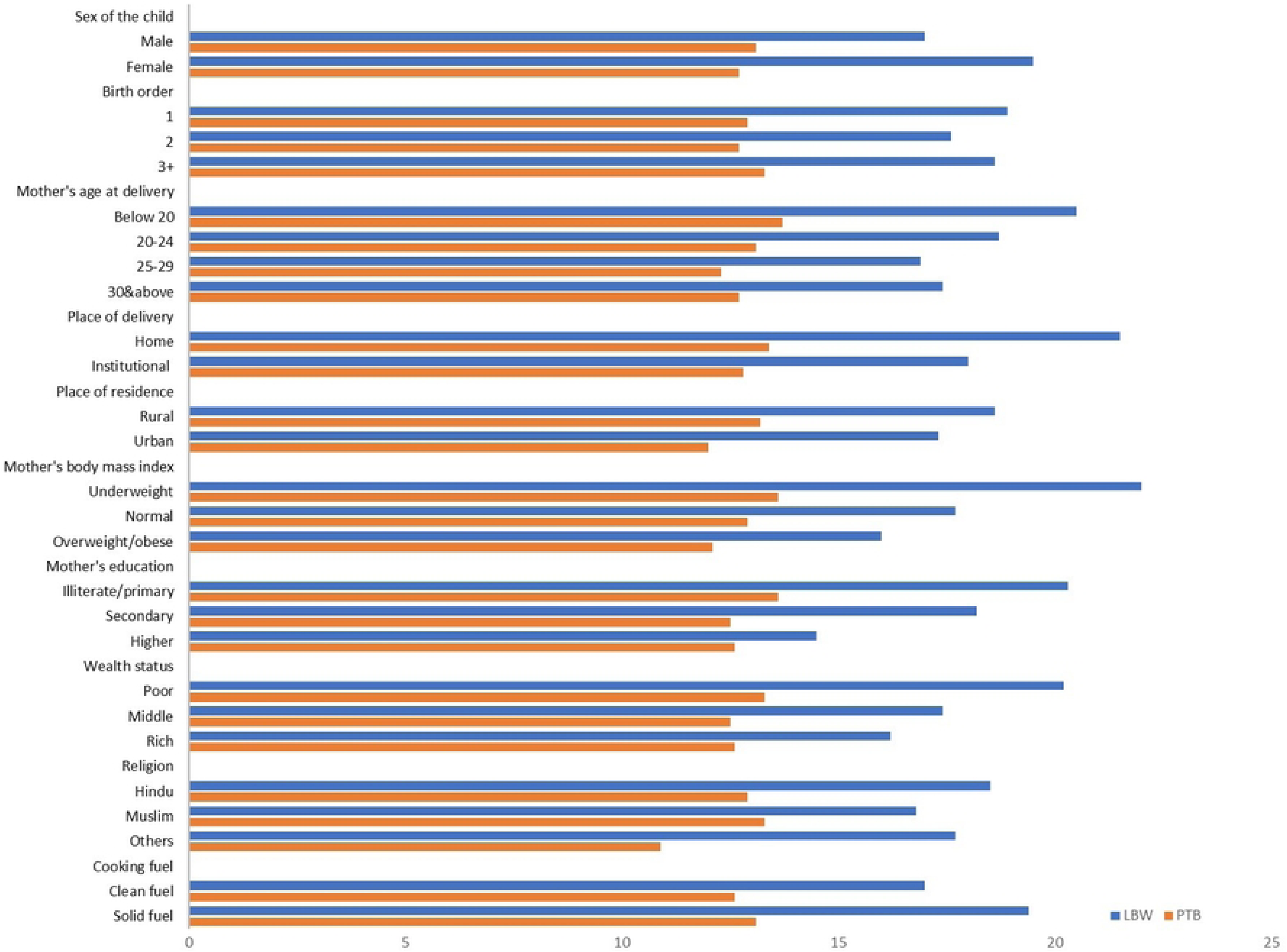
Weighted percentage of low birth weight and preterm birth by background characteristics

Our unadjusted and adjusted multivariate logistic regression models consistently revealed associations between air pollution during pregnancy and birth outcomes, as shown in **Table 2**. Higher ambient PM2.5 concentrations during pregnancy were associated with higher odds of both low birth weight (LBW) (AOR: 1.37; 95% CI: 1.29– 1.45) and preterm birth (PTB) (AOR: 1.67; 95% CI: 1.57–1.77). The odds ratios were higher in the unadjusted models for both LBW (OR: 1.56; 95% CI: 1.50–1.63) and PTB (OR: 1.62; 95% CI: 1.55–1.69). A slight increase in temperature was associated with higher odds of LBW (AOR: 1.03; 95% CI: 1.01–1.04), though it was not significantly associated with PTB, while higher rainfall was significantly associated with both LBW (AOR: 1.07; 95% CI: 1.03–1.12) and PTB (AOR: 1.04; 95% CI: 1.02–0.10). The use of solid fuel for cooking was associated with higher odds of LBW (AOR: 1.04; 95% CI: 1.01–1.07), but it was not significantly associated with PTB. Female children had higher odds of being born with LBW (AOR: 1.18; 95% CI: 1.15– 1.20) compared to male children but had slightly lower odds of PTB (AOR: 0.97; 95% CI: 0.95–1.00). Higher birth order was associated with lower odds of LBW (AOR for second birth: 0.89; 95% CI: 0.86–0.91; AOR for third or higher birth: 0.84; 95% CI: 0.80–0.88). For PTB, only a third or higher birth order showed a significant decrease in odds (AOR: 0.94; 95% CI: 0.89–0.98). Teenage mothers (below 20 years) had higher odds of giving birth to children with LBW (AOR: 1.09; 95% CI: 1.04–1.15) and PTB (AOR: 1.08; 95% CI: 1.02–1.14).

**Table 2.**
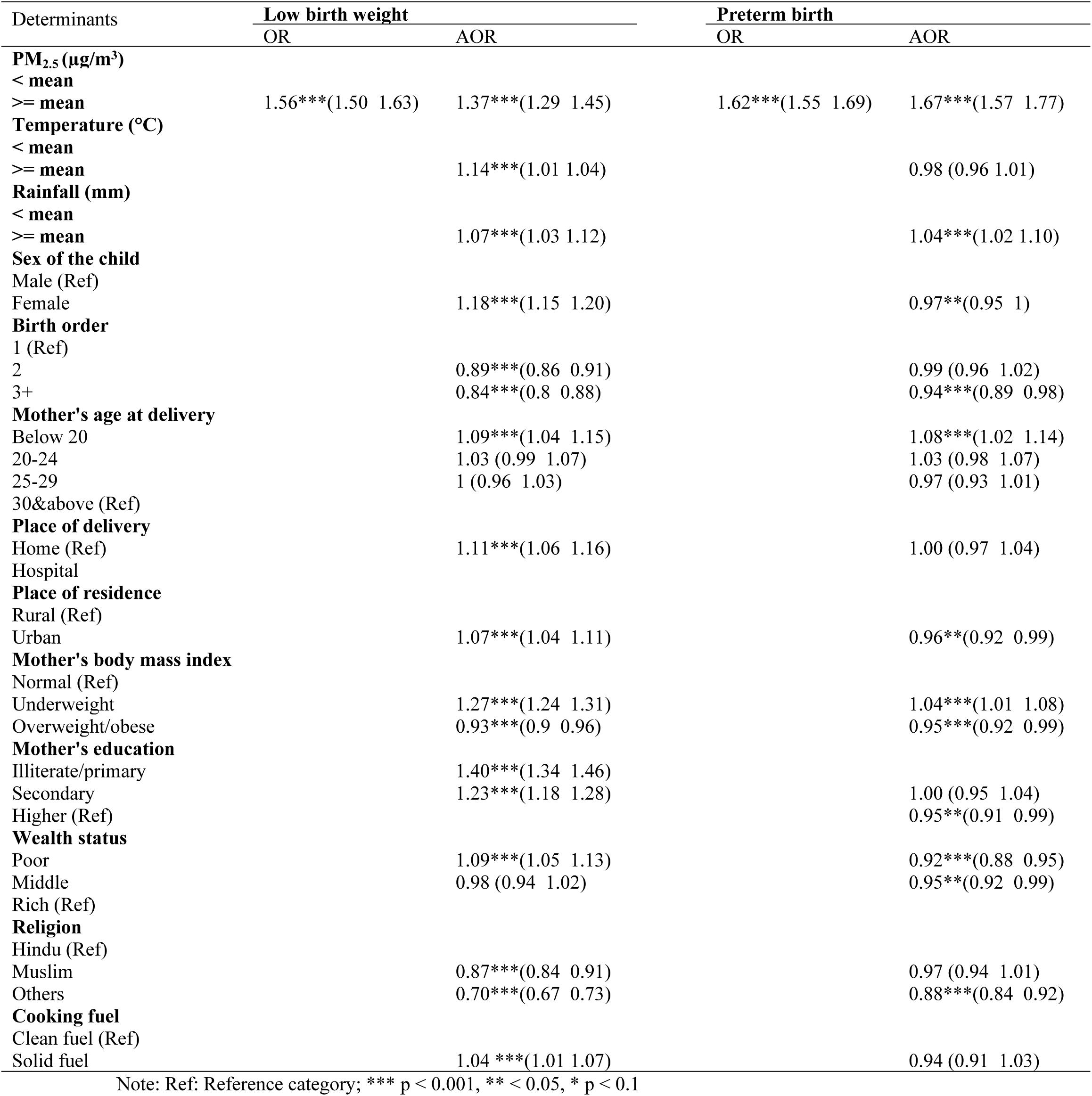
Adjusted and unadjusted odds ratio showing the effects of in-utero exposure to PM_2.5_ on low birth weight and preterm birth.

Children born at home had higher odds of LBW (AOR: 1.11; 95% CI: 1.06–1.16). Urban residence was associated with higher odds of LBW (AOR: 1.07; 95% CI: 1.04–1.11) and lower odds of PTB (AOR: 0.96; 95% CI: 0.92–0.99). Underweight mothers had higher odds of both LBW (AOR: 1.27; 95% CI: 1.24–1.31) and PTB (AOR: 1.04; 95% CI: 1.01–1.08), while overweight or obese mothers had lower odds of both LBW (AOR: 0.93; 95% CI: 0.90–0.96) and PTB (AOR: 0.95; 95% CI: 0.92–0.99). Lower maternal education levels were associated with higher odds of LBW (AOR for illiterate/primary: 1.40; 95% CI: 1.34–1.46; AOR for secondary: 1.23; 95% CI: 1.18–1.28), while higher education was associated with lower odds of PTB (AOR: 0.95; 95% CI: 0.91–0.99). Poor mothers had higher odds of LBW (AOR: 1.09; 95% CI: 1.05–1.13) and lower odds of PTB (AOR: 0.92; 95% CI: 0.88–0.95). Muslim mothers had lower odds of LBW (AOR: 0.87; 95% CI: 0.84–0.91) and similar odds of PTB compared to Hindu mothers.

There was a high likelihood of LBW in women who experienced high levels of PM_2.5_ during their gestational period (Figure 3 **a and 2b**). Employing a distributed spline approach, the study identified a growing trend in the risk of delivering LBW babies as the exposure level increased, especially after the exposure level of 40 PM_2.5_ ug/m^3^. Concerning PTB, the odds displayed a rapid and exponential increase in relation to the PM_2.5_ exposures and the increasing trend became notably rapid after reaching the exposure level of 50 PM_2.5_ ug/m^3^.

**Figure 3.**
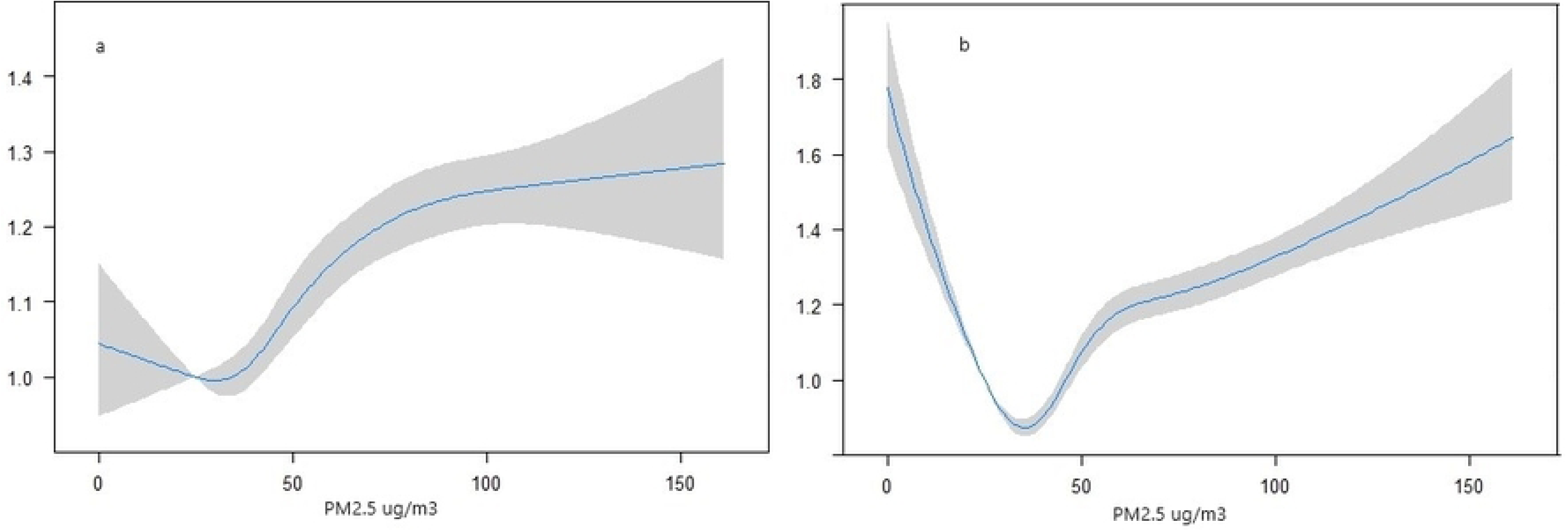
Susceptibility to (a) low birth weight, and (b) preterm birth due to the in-utero exposure to PM2.5.

### 3.4 Sensitivity analysis

The unadjusted odds ratio demonstrates a significantly higher likelihood of LBW associated with PM2.5 exposures during pregnancy (OR: 1.56; 95% CI: 1.50–1.63). However, the strength of this association decreased after adjusting for environmental, socioeconomic, maternal, and child characteristics (**Table 3**). In contrast, for PTB, the odds ratio shifted from 1.62 in the unadjusted model to 1.67 after accounting for these determinants. This discrepancy underscores the influence of the various determinants considered in the study on the odds ratio values, highlighting their importance in the analysis.

**Table 3.**
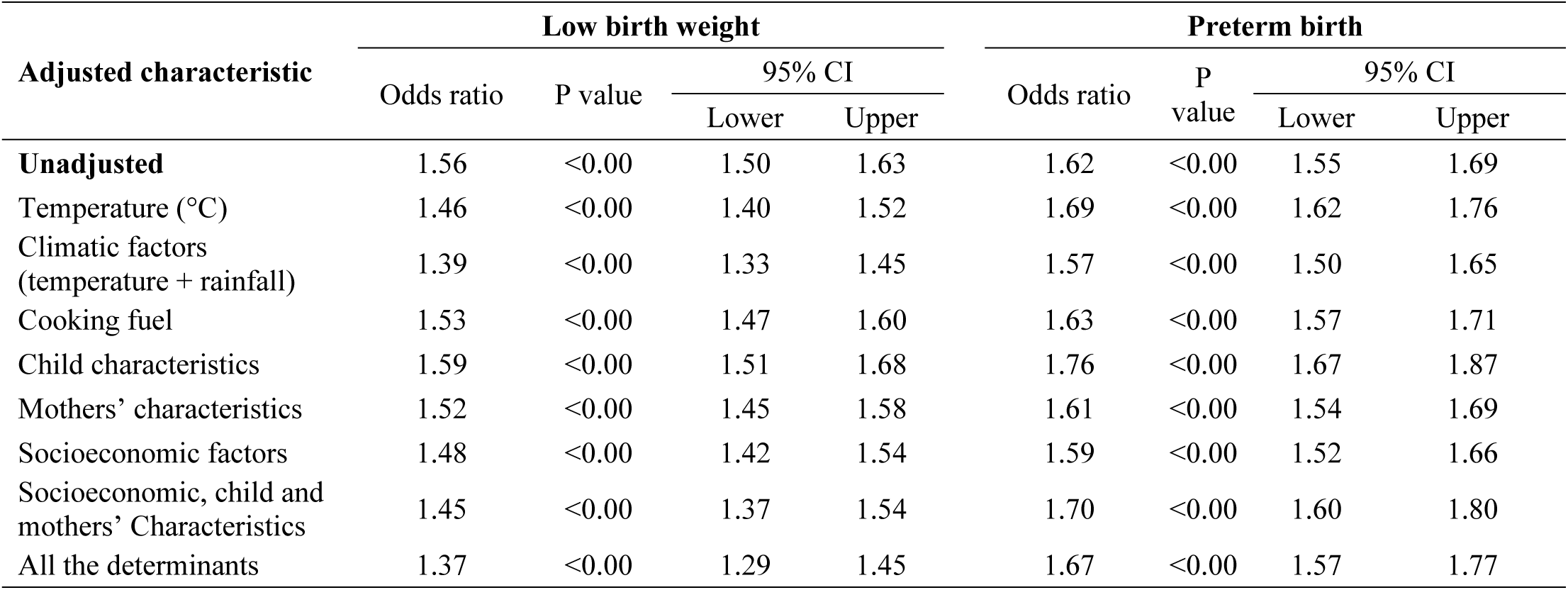
Sensitivity analysis of the effects of PM_2.5_ on low birth weight and preterm birth.

### 3.5 Spatial association between PM_2.5_ and ABO

The findings of the bivariate LISA map showed that children living in the Northern districts of India faced a higher vulnerability to ambient air pollution as the high-high clusters of spatial association were found in Punjab, Delhi, Madhya Pradesh, Rajasthan and some parts of Uttar Pradesh (Figure 4). A total of 109 districts had a significant association between in-utero exposure to PM_2.5_ and LBW. On the other hand, 40 districts had high-high clusters of autocorrelations between PM_2.5_ and PTB. Notably, it was observed that most of the districts of Uttar Pradesh were found to be more vulnerable to PM_2.5_ in the context of preterm birth.

**Figure 4.**
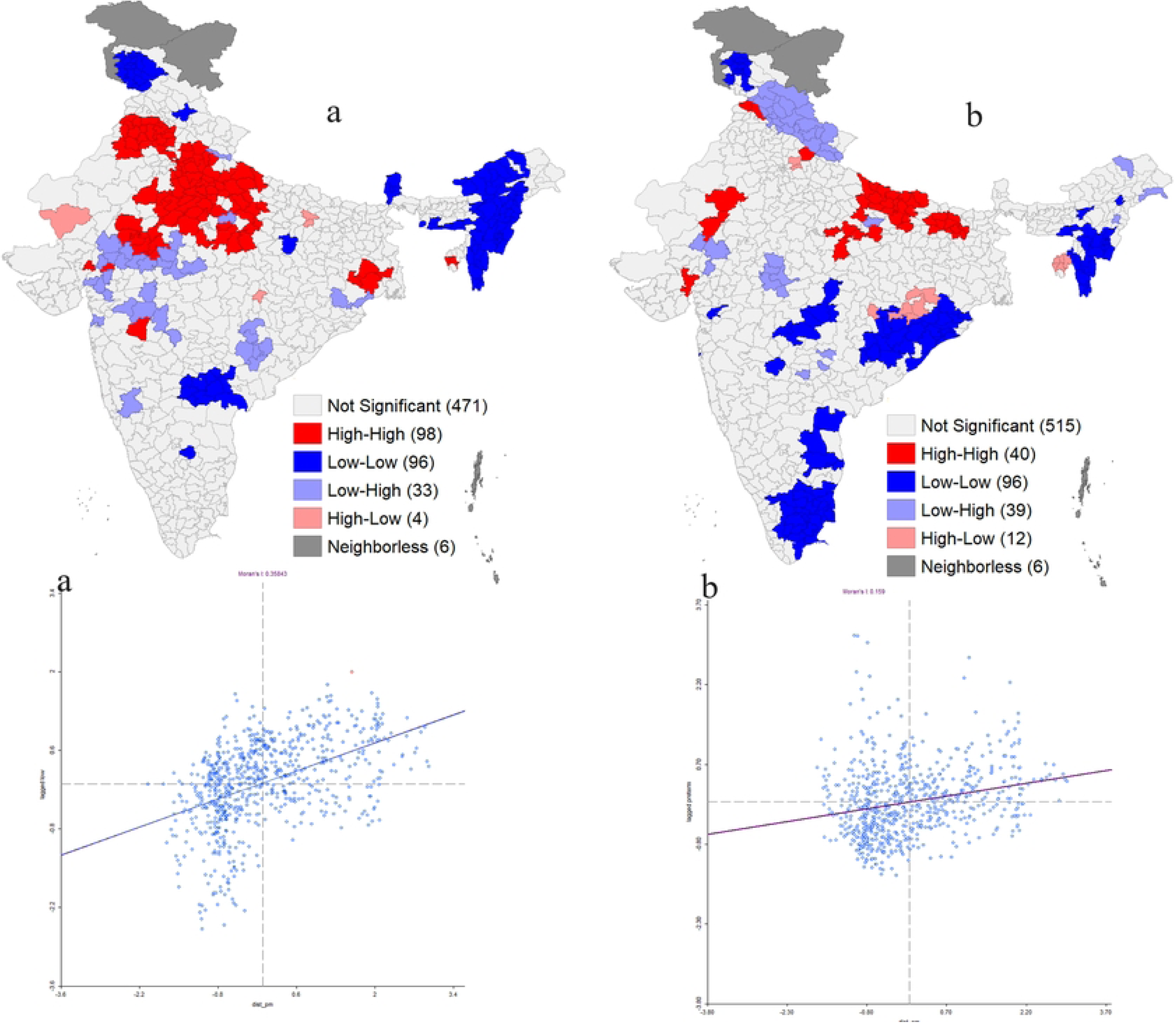
Bivariate LISA map showing the spatial association between in-utero exposure to PM_2.5_ and low birth weight (a) and preterm birth (b).

Additionally, the study employed OLS, GWR, MGWR, SLM, and SEM, to investigate the spatial relationship between PM_2.5_ exposures during pregnancy and the occurrences of LBW and PTB, while considering potential confounding factors. In line with our hypothesis, the analysis revealed a robust spatial association between PM_2.5_ exposures and LBW, although the strength of this association varied across the different models. Notably, the GWR model showed the highest impact of PM_2.5_ exposures (β=0.122, SE=0.009), while MGWR demonstrated the highest goodness-of-fit (R^2^ = 68%) (**Table 4**). Regarding PTB, the MGWR model provided the best explanatory power, accounting for 54% of the variance, surpassing other models. In the GWR model, a noteworthy finding was that a one-unit increase in PM_2.5_ exposure corresponded to a 5% increase in the prevalence of PTB (**Table 5**) and a 12% increase in LBW. Overall, a reduction in the prevalence of LBW and PTB was observed across all models, with MGWR and GWR models showing particularly promising outcomes. Nevertheless, higher prevalence rates persisted in Northern India and certain parts of the eastern region, most notably in Odisha (Figure 5).

**Figure 5.**
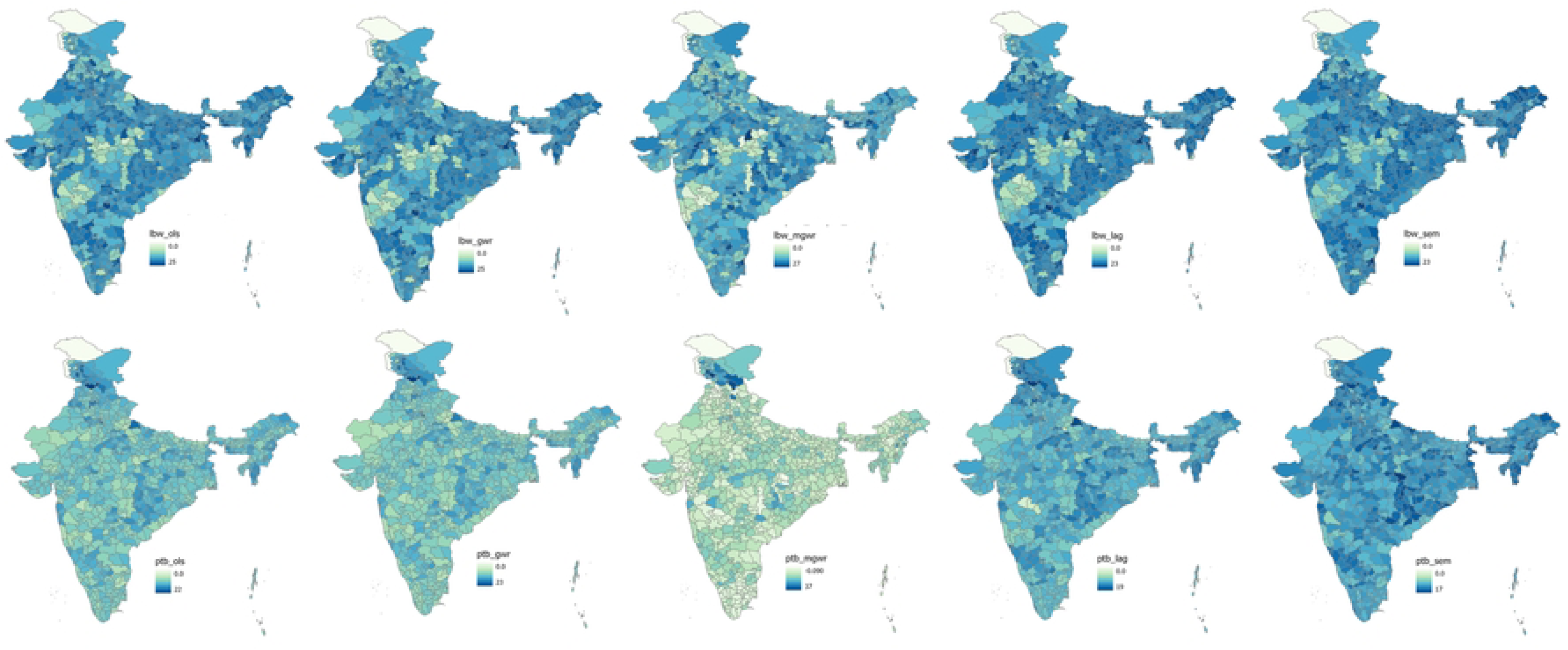
Predicted low birth weight (LBW) and preterm birth (PTB) results from Ordinary List Square (OLS), Geographically Weighed Regression (GWR), Multiscale Geographically Weighed Regression (MGWR), Spatial Lag Model (SLM) and Spatial Error Model (SEM) after adjusting environmental, Socioeconomic maternal and child characteristics.

**Table 4.** spatial regression models showing the spatial association between PM_2.5_ and low birth weight adjusted confounding factors.

| Variables | OLS |  | GWR |  | MGWR |  | SLM |  | SEM |  |
| --- | --- | --- | --- | --- | --- | --- | --- | --- | --- | --- |
|  | Coefficient | SE | Coefficient | SE | Coefficient | SE | Coefficient | SE | Coefficient | SE |
| PM<sub>2.5</sub> (µg/m<sup>3</sup>) | 0.059 | 0.009 | 0.122 | 0.009 | 0.119 | 0.016 | 0.043 | 0.014 | 0.031 | 0.008 |
| Underweighted mother | 0.159 | 0.031 | 0.179 | 0.007 | 0.163 | 0.003 | 0.124 | 0.032 | 0.102 | 0.027 |
| Illiterate mother | 0.061 | 0.018 | 0.095 | 0.009 | 0.013 | 0.001 | 0.068 | 0.021 | 0.040 | 0.016 |
| Belong to poor family | -0.053 | 0.015 | 0.234 | 0.008 | 0.494 | 0.000 | -0.011 | 0.019 | -0.022 | 0.013 |
| Hindu religion | 0.010 | 0.009 | -0.055 | 0.006 | -0.017 | 0.002 | 0.007 | 0.011 | -0.002 | 0.008 |
| Non-institutional delivery | -0.083 | 0.025 | -0.067 | 0.008 | -0.155 | 0.000 | -0.060 | 0.027 | -0.057 | 0.022 |
| Using solid cooking fuel | 0.062 | 0.014 | 0.126 | 0.009 | -0.118 | 0.000 | 0.024 | 0.017 | 0.028 | 0.013 |
| Belong to rural areas | -0.018 | 0.012 | -0.137 | 0.008 | -0.114 | 0.005 | 0.002 | 0.012 | -0.001 | 0.011 |
| Female child | -0.005 | 0.058 | 0.003 | 0.003 | 0.001 | 0.004 | 0.008 | 0.048 | 0.001 | 0.050 |
| Teen aged mothers | 0.022 | 0.034 | 0.066 | 0.007 | 0.101 | 0.007 | 0.023 | 0.040 | 0.005 | 0.030 |
| Having 3& more births | -0.090 | 0.046 | -0.122 | 0.007 | -0.03 | 0.005 | -0.102 | 0.048 | -0.069 | 0.040 |
| Average rainfall (°C) | 0.000 | 0.003 | -0.071 | 0.005 | -0.055 | 0.000 | -0.003 | 0.003 | 0.001 | 0.003 |
| Average temperature (mm) | 0.041 | 0.041 | 0.026 | 0.007 | 0.165 | 0.000 | 0.105 | 0.052 | 0.023 | 0.036 |
| R<sup>2</sup> | 0.34 |  |  |  | 0.68 |  | 0.49 |  | 0.52 |  |
| AIC | 4192.36 |  |  |  | 1420.83 |  | 4045.78 |  | 4022.85 |  |

**Table 5.** spatial regression models showing the spatial association between PM_2.5_ and preterm birth adjusted confounding factors.

| Variables | OLS |  | GWR |  | MGWR |  | SLM |  | SEM |  |
| --- | --- | --- | --- | --- | --- | --- | --- | --- | --- | --- |
|  | Coefficient | SE | Coefficient | SE | Coefficient | SE | Coefficient | SE | Coefficient | SE |
| PM<sub>2.5</sub> (µg/m<sup>3</sup>) | 0.043 | 0.016 | 0.053 | 0.012 | 0.041 | 0.018 | 0.018 | 0.014 | 0.035 | 0.024 |
| Underweighted mother | 0.009 | 0.053 | 0.051 | 0.014 | 0.044 | 0.012 | -0.022 | 0.046 | -0.059 | 0.056 |
| Illiterate mother | 0.035 | 0.030 | 0.169 | 0.011 | 0.214 | 0.000 | 0.030 | 0.026 | 0.070 | 0.036 |
| Belong to poor family | -0.055 | 0.026 | -0.121 | 0.018 | -0.140 | 0.000 | -0.024 | 0.022 | -0.029 | 0.033 |
| Hindu religion | 0.050 | 0.016 | 0.078 | 0.010 | 0.038 | 0.003 | 0.023 | 0.014 | 0.023 | 0.019 |
| Non-institutional delivery | 0.050 | 0.043 | 0.047 | 0.019 | 0.071 | 0.006 | 0.022 | 0.037 | 0.008 | 0.046 |
| Using solid cooking fuel | -0.014 | 0.025 | -0.096 | 0.015 | -0.045 | 0.005 | -0.019 | 0.021 | -0.048 | 0.030 |
| Belong to rural areas | 0.044 | 0.021 | 0.054 | 0.005 | 0.062 | 0.000 | 0.027 | 0.018 | 0.042 | 0.021 |
| Female child | 0.032 | 0.100 | 0.031 | 0.009 | 0.045 | 0.004 | 0.030 | 0.086 | 0.038 | 0.084 |
| Teen aged mothers | 0.028 | 0.059 | 0.022 | 0.005 | 0.022 | 0.001 | 0.019 | 0.051 | 0.027 | 0.069 |
| Having 3& more births | 0.053 | 0.078 | 0.096 | 0.009 | 0.05 | 0.013 | 0.014 | 0.068 | 0.003 | 0.082 |
| Average rainfall (°C) | 0.000 | 0.005 | 0.030 | 0.008 | 0.030 | 0.003 | 0.000 | 0.004 | -0.001 | 0.005 |
| Average temperature (mm) | -0.291 | 0.070 | -0.143 | 0.013 | -0.112 | 0.009 | -0.117 | 0.061 | -0.106 | 0.089 |
| R<sup>2</sup> | 0.17 |  |  |  | 0.54 |  | 0.29 |  | 0.31 |  |
| AIC | 4953.57 |  |  |  | 1645.21 |  | 4810.00 |  | 4796.55 |  |

## 4. Discussion

The study is the first attempt to measure preterm birth at the Indian district level by exploring the calendar data of NFHS ^62^. The analysis establishes the evidence of the association between in-utero exposure to PM_2.5_ and adverse birth outcomes by leveraging satellite data and large-scale survey data. The individual-level analysis reveals that an increase in ambient PM_2.5_ is associated with a greater likelihood of LBW and PTB, consistent with previous studies ^36,67–69^. We observe a higher value of the odds ratio of having preterm infants whose mothers were exposed to PM_2.5_ during pregnancy as compared to the results for LBW. A significant role of indoor air pollution has been observed in the study for LBW. Further, climatic factors such as rainfall and temperature are significantly associated with ABOs in India. To the best of our knowledge, this study is the first of its kind to understand the association between in-utero exposure to PM_2.5_ and adverse birth outcomes in India.

It is worth noting that except for Nagaland, North-Eastern states had a lower prevalence of preterm birth and low birth weight, coinciding with lower pollution levels in that region. Previous studies have found that the concentration of ambient PM_2.5_ is highest in the states of upper-Gangetic plains such as Delhi, Punjab, Haryana, Uttar Pradesh and Bihar, as observed in the present study ^70–73^. A recent Lancet study suggests that the average PM_2.5_ concentrations in Delhi were 13.8 times higher than that in Kerala ^74^. The Northern states had the highest PM_2.5_ concentrations. The NFHS-5 report suggests that a large proportion of households in the Northern parts of India use solid fuels compared to other regions ^46^. It is well documented that the residential sector is a significant contributor to the total PM_2.5_ emissions along with the industry, energy and agriculture sectors ^75^. Among industrial, residential and energy sources, the contribution of energy sources to total emissions is the maximum, while residential sources contribute the maximum to PM_2.5_ emissions during winter and post-monsoon ^75^. In contrast, certain studies conducted on future emissions scenarios in India forecast an increase in PM_2.5_ levels ^76^. However, at the urban or city level, where most households are already using cleaner fuel, reducing vehicular emissions (both exhaust and non-exhaust) emerges as a crucial strategy for reducing PM_2.5_ levels. It was prominently observed during the coronavirus pandemic (COVID-19) lockdown in Indian cities when traffic reduction substantially minimized urban areas’ exposure to air pollutants ^70,77^. Moreover, the issue of air pollution is exacerbated by crop residue burning and forest fires in northern India, significantly contributing to the toxic air quality in the region ^78,79^. The higher prevalence of low birth weight and preterm birth in the districts of the Northern region indicates a spatial association between PM_2.5_ and birth outcomes. In line with our hypothesis, the study finds an individual level and spatial association between in-utero exposure to PM_2.5_ and LBW and PTB.

The mechanisms behind preterm birth due to exposure to PM_2.5_ are not clearly understood. Some epidemiological and toxicological studies proposed different pathways to explain the paradox. Due to the finer size of PM_2.5,_ inhaling particulate matter deposits in the lungs and affects the circulatory system ^80^. Which is a reason of having oxidative stress, blood coagulation and placental inflammation that restricts the fetal growth ^81–85^. Nevertheless, the presence of particulate matter in the human body disturbs oxygen transport and causes hormone dysfunction, which is the reason for placental insufficiency ^86–89^.

An earlier study identified that maternal exposure to PM_2.5_ in the latter stages of pregnancy induces cytokine activation, favouring inflammation ^90^. Sometimes inhaled PM_2.5_ penetrates the toxic gases that damage deoxyribonucleic acid (DNA), restricting nutrient supply to the fetus resulting in pregnancy complications during the last trimester of pregnancy that triggers the possibility of having a preterm birth ^91,92^. Moreover, substantial exposure to a large amount of PM_2.5_ has been linked to fetal malformation, miscarriage and stillbirth, all of which can influence subsequent birth outcomes ^93–95^.

Medical studies found that the mother’s fetus grows rapidly in the third trimester of pregnancy ^96,97^. That period is more sensitive, and exposure to PM_2.5_ can hemorrhage the foetal growth easily ^98,99^. Nevertheless, the level of thyroid hormone might be affected by the toxicity of PM_2.5_, which is a responsible factor for less fetus weight ^100^. Previous studies have found that fetal growth depends on many factors, such as mothers’ health, socioeconomic condition and genetic factors ^101,102,102,103^. Thus, the present study has adjusted for those potential plausible factors. Nevertheless, the study establishes a significant association between PM_2.5_ and LBW. Further investigation is required to explore the biological mechanism by considering the biomarker measurement of the fetus.

Low birth weight and preterm birth significantly contribute to the highest number of stunting and premature deaths in India ^104–106^. Since 1970, the Indian government has been trying to improve child health by adopting several schemes that promote the utilization of maternal healthcare facilities, spread awareness of reproductive health, supply nutrients to mothers and newborns, etc. Adverse birth outcomes not only impact a child’s health but also reduce the productivity of the human resources of a country.

Understanding the impact of exposure to indoor air pollution during pregnancy is also important because pregnant women tend to spend most of their time indoors and this time only increases once the pregnancy progresses. Past studies found that releasing pollutants from uncleaned biomass burning in the household restricts fetal growth, increasing the probability of having a child with low weight and preterm birth ^107,108^. In line with the hypothesis, our study also found a negative association between in-utero exposure to PM_2.5_ and birth weight, whereas the result for PTB was the opposite. As PTB mostly depends on the mother’s health and obstetric factors ^62^, which could explain the absence of a discernible effect of PM_2.5_ on PTB in the study. However, addressing indoor air pollution through interventions such as promoting cleaner fuels and improving home ventilation is crucial for improving pregnancy outcomes and neonatal health.

The present finds a significant role of climatic factors for ABOs. Previous evidence suggests that high temperatures can cause heat stress, dehydration, and reduced uteroplacental blood flow, impairing fetal growth and increasing the risk of LBW ^109^. Additionally, heat-induced oxidative stress and inflammation can disrupt placental function, while cardiovascular strain and hormonal imbalances can further complicate pregnancy outcomes. On the other hand, extreme rainfall can heighten the risk of waterborne and vector-borne diseases, leading to maternal infections that adversely affect fetal development (Poursafa et al., 2015). Flooding can also result in nutritional deficiencies, physical and psychological stress, and disrupted healthcare access, all of which contribute to LBW and PTB ^110,111^. Improved healthcare access, infrastructure development, and policies addressing climate change are essential to protect maternal and neonatal health against these challenges.

Our findings suggest that both indoor and ambient air pollution along with climatic factor are significantly associated with ABOs. However, to gain a deeper understanding of the biological mechanisms underpinning the relationship between air pollution and malnutrition, which cannot be fully explored using existing datasets, conducting an in-depth epidemiological study is imperative. It is important to note that our research assumes that mothers did not change their residence during the period from pregnancy to the conducting date of the survey. The study acknowledges that like some other possible determinants, smoking practices and alcohol consumption were not considered in the model due to the under-reporting nature. Also, the possibility of biases due to self-reporting cannot be ignored. Due to the cross-sectional nature of the study, we cannot draw a causal relationship, other aspects including the seasonality effects can be explored using the longitudinal study in the future. As India is not satisfactorily progressing in child nutrition, hence consorted effort to improve environmental conditions, especially air quality, is essential to tackle issues related to child health.

## 5. Conclusion

The present study provides robust evidence linking in-utero exposure to ambient PM_2.5_ and climatic factors, such as temperature and rainfall, with adverse birth outcomes in India. The geostatistical analysis underscores the need for targeted interventions, particularly in Northern districts identified as highly vulnerable. To address these challenges, comprehensive strategies are essential. The National Clean Air Program should be intensified, with stricter emission standards and enhanced air quality monitoring. Integrating air quality data with health surveillance systems will enable precise identification of at-risk populations. Additionally, policies promoting clean cooking fuels and energy-efficient technologies can reduce indoor air pollution. Climate adaptation strategies, such as developing heat action plans and improving water management, should be incorporated into public health planning to mitigate the effects of extreme temperatures and irregular rainfall. Public health initiatives must raise awareness about the risks of air pollution and climate change, particularly among pregnant women.

## Statements & Declarations

### Data Availability

The study uses secondary data that are available on reasonable request through https://dhsprogram.com/data/dataset_admin/

### Ethics approval

The Indian Demographic and Health Survey (DHS) is known as National Family Health Survey (NFHS) in India. We used published large scale national data where every respondent was anonymized in the data set itself. As it is not based on a primary survey-cases, we need not to do any anonymization in the study as the data is already made in that fashion following all ethical protocols. Thereby, it is certified that all applicable institutional and governmental regulations concerning the ethical use of human volunteers were followed during the course of the survey.

### Consent to participate

Verbal as well as written informed consent was obtained from all the participants. The informed consent was taken from their parent or legal guardian who were not mature or below 18 aged. Then blood sample was taken from the finger and collected in microcuvette. The data is in the public domain-free of cost for users and funded by the government. More details on that available at https://dhsprogram.com/search/index.cfm?bydoctype=publication&bypubtype=5.

### Consent to publish

The dataset is publicly available, thus consent for publication is not applicable for the study.

